# “Salivary dim-light melatonin onset in early Amyotrophic Lateral Sclerosis predicts functional decline, respiratory symptom emergence, and survival”

**DOI:** 10.64898/2026.04.24.26351642

**Authors:** Alessandro Bombaci, Antonella Iadarola, Alessia Giraudo, Elisa Fattori, Serena Sinagra, Alice Magnino, Andrea Calvo, Adriano Chiò, Alessandro Cicolin

**Affiliations:** Vita-Salute San Raffaele University, Milan, Italy; IRCCS Policlinico San Donato, San Donato Milanese, Italy; Turin ALS Centre, “Rita Levi Montalcini” Department of Neuroscience, University of Torino, Turin, Italy; University of Genova, Department of Neuroscience, Rehabilitation, Ophthalmology, Genetics, Child and Maternal Health (DINOGMI), Italy; Sleep Medicine Centre, “Rita Levi Montalcini” Department of Neuroscience, University of Torino, Turin, Italy

**Keywords:** melatonin, neurodegeneration, biomarker, DLMO, sleep, circadian

## Abstract

**Background:** Sleep–wake and circadian disturbances are increasingly recognised in people living with amyotrophic lateral sclerosis (plwALS), but endogenous circadian phase timing and its prognostic significance in early disease remain unclear. We assessed whether salivary dim-light melatonin onset (DLMO), an objective marker of central circadian phase, is altered in early plwALS and whether it provides prognostic information.

**Methods:** In this prospective longitudinal observational study, plwALS within 18 months of symptom onset underwent home-based salivary melatonin sampling under dim-light conditions at six predefined time points around habitual sleep onset (HSO). Melatonin profiles were modeled using cubic smoothing splines, and DLMO was defined as the first time the fitted curve reached 3 pg/mL. Clinical, respiratory, and sleep assessments were collected at baseline (T0) and after 6 months (T6); a subgroup repeated saliva sampling at T6. Age- and sex-matched controls underwent melatonin profiling. Associations with disease progression, incident respiratory symptoms, and survival/tracheostomy were examined using regressions and survival analyses.

**Results:** Fifty plwALS were enrolled. Compared with controls, plwALS showed an earlier DLMO (20:24±1:18 vs 20:58±0:50; p=0.028) despite similar HSO and chronotype. Within ALS cohort, a later baseline DLMO correlated with worse functional/motor status, faster progression of disease, incident dyspnea/orthopnea by T6 (adjusted OR 3.02; p=0.017), and poorer survival/tracheostomy-free outcome. In re-sampled subgroup (n=28), DLMO and other melatonin-derived metrics did not change over ∼6 months.

**Conclusions:** Circadian phase alterations are detectable in early-ALS. Baseline DLMO may represent a non-invasive prognostic biomarker for progression, respiratory symptom emergence and survival, warranting validation in larger multicentre cohorts.

**Key messages:** *What is already known on this topic:* Sleep and circadian disturbances are increasingly recognised as early, biologically relevant non-motor features of amyotrophic lateral sclerosis (ALS), and recent translational and neuroimaging studies support early involvement of sleep-regulatory and hypothalamic networks. Dim-light melatonin onset (DLMO) is an established objective marker of central circadian phase, but endogenous melatonin timing in ALS and its prognostic relevance have not been previously defined.

*What this study adds:* In a prospective cohort of patients with early-ALS, salivary DLMO was altered relative to matched controls, and within the ALS cohort a later baseline DLMO was associated with worse functional and motor status, faster subsequent progression, incident respiratory symptoms at 6 months, and poorer survival/tracheostomy-free outcome. These findings identify circadian phase timing as a clinically informative signal in early-ALS.

*How this study might affect research, practice or policy:* If validated, DLMO could complement established prognostic tools in early-ALS and support enrichment of phase-aware clinical trials. They also provide a rationale for phase-aware longitudinal studies integrating circadian phenotyping, respiratory, imaging, and plasmatic biomarker and for testing whether interventions targeting circadian alignment can improve symptoms or clinical trajectories in selected patients with ALS.

## Introduction

Amyotrophic lateral sclerosis (ALS), the most common form of motor neuron disease, is characterized by progressive degeneration of both upper and lower motor neurons, leading to bulbar, limb, truncal and respiratory weakness^1^. Increasingly, ALS is viewed as a multisystem disorder in which cognitive-behavioural, autonomic, metabolic, sensory, and sleep-related symptoms contribute substantially to disease burden^2–4^. This broader perspective has increased interest in accessible, biologically informative, and clinically useful biomarkers across early disease stages. Alongside neurofilaments^5,6^ and other emerging fluid markers^7^, there remains a need for noninvasive measures that capture distinct aspect of disease biology and help refine prognostic stratification.

Sleep and circadian disturbances are common in ALS and have classically been attributed to nocturnal hypoventilation, cramps, pain, immobility, or sleep-disordered breathing^8^. However, recent work suggests sleep abnormalities may arise early in ALS, including in presymptomatic mutation carriers, and may therefore reflect intrinsic network dysfunction rather than only late mechanical or respiratory complications^9,10^. In parallel, neuroimaging studies indicate hypothalamic involvement across the ALS-frontotemporal spectrum, supporting the biological plausibility of circadian dysregulation^11^.

Melatonin (MLT; N-acetyl-5-methoxytryptamine) has emerged as a candidate biomarker and therapeutic agent in neurodegeneration. Secreted by the pineal gland under control of the suprachiasmatic nucleus, melatonin regulates circadian phase and sleep initiation and can be quantified in saliva with ambulatory protocols^12,13^. The dim-light MLT onset (DLMO), the time at which MLT levels rises above a predefined threshold under dim conditions, is the accepted gold-standard phase marker of the human circadian clock^12–14^. At-home salivary DLMO sampling is increasingly standardized, but its validity still depends on careful control of light exposure, sampling timing, and analytic methodology^12,14^.

Across neurodegenerative disorders, altered MLT dynamics have been linked to sleep–wake and cognitive symptoms. In Parkinson’s disease, impaired MLT rhythmicity has been linked to excessive daytime sleepiness and broader circadian dysfunction, and dopaminergic therapy may advance MLT phase^15^. In Alzheimer’s disease, night-time MLT levels are reduced and delayed even in mild-to-moderate stages^15,16^, while exogenous MLT has shown mixed and overall modest clinical benefits, highlighting the need for standardized dosing and phenotyping protocols^17^. In ALS specifically, endogenous MLT profiling remains scarce. Yet preclinical studies show that MLT can delay disease progression and mortality in SOD1 models and modulate autophagy- and apoptosis-related pathways^18,19^ while observational human data have suggested a possible association between MLT use and slower progression^20^. However, these findings do not establish whether endogenous circadian timing is altered in people living with ALS (plwALS) or whether such alterations carry prognostic information.

More broadly, MLT’s combined chronobiotic and cytoprotective actions (including antioxidant, anti-inflammatory, mitochondrial-stabilizing, and potential facilitation of glymphatic clearance) make it a biologically plausible node within ALS pathobiology, in line with clinical perspectives in neurodegeneration^21^.

Accordingly, we prospectively evaluated salivary MLT secretion in early-stage plwALS using home-based dim-light sampling, with particular focus on DLMO as a marker of central circadian phase. We aimed to determine whether MLT timing is altered relative to healthy controls (HCs) and whether phase-based metrics are associated with clinical status, respiratory symptom emergence, disease progression, and survival.

## 2. Materials and Methods

### 2.1 Study Design and participants

This prospective longitudinal observational study was conducted between August 2021 and August 2023 at Sleep Centre and at ALS Expert Centre, Molinette Hospital, University of Turin, Italy. plwALS were diagnosed according to the Gold Coast criteria^22^. We enrolled plwALS within 18 months of symptom onset with non-advanced disease, defined as ALS Functional Rating Scale-revised (ALSFRSr)^23^ total score >36 and a bulbar subscore >6 (ALSFRSr_B, calculated as the sum of items 1 and 3), who had received riluzole for at least 2 months, and were able to provide informed consent. Age- and sex-matched HCs without history of sleep or neurological disorders were also included.

All procedures adhered to Good Clinical Practice and local ethics approval (protocol number: 0028226 of 15/Mar/2021), ensuring anonymization of samples and data confidentiality.

Exclusion criteria for plwALS and HCs comprised: chronic respiratory or sleep disorders within six months of screening, forced vital capacity (FVC) <60% of predicted, abnormal screening questionnaires for insomnia, restless legs syndrome (RLS), or risk of obstructive sleep apnoea (OSA), exogenous MLT therapy, medications known to affect MLT secretion (e.g. beta-blockers, lithium, thyroid hormone replacement, antidepressants, benzodiazepines, antipsychotics), active cancer, infections, and autoimmune disorders.

### 2.2 Clinical assessment

At baseline (T0), all patients underwent clinical assessment including demographic characteristics, smoking status, caffeine intake, and a neurological examination (including the Medical Research Council scale [MRC]^24^ assessment and ALSFRSr scale). Validated sleep questionnaires (Insomnia Severity Index score [ISI], STOP-Bang questionnaire, RLS questionnaire, and Morningness-Eveningness Questionnaire [MEQ], respectively useful for the evaluation of insomnia, of OSA, of RLS, and of chronotype), polysomnography (PSG), spirometry (including evaluation of FVC, Maximum Expiratory Flow at 50% [MEF50], and Peak Expiratory Flow [PEF]), and arterial blood gas analysis (ABG) early in the morning were performed at T0. The same clinical, respiratory, and sleep assessments were repeated after approximately 6 months (T6).

Saliva samples were collected from plwALS at T0 and, in a subgroup of 28 plwALS, again at T6. All 48 HCs underwent neurological examination and PSG; 15 also provided saliva samples.

ALSFRSr subscores used included: ALSFRSr_noresp (excluding respiratory items), ALSFRSr_B, and ALSFRSr_4limb (limb score: items 4–9).

Electromyography, cerebral MRI with tractography, neuropsychological evaluation^25^, and genetic testing for the most common mutation (*C9orf72, SOD1, TARDBP and FUS* genes) were performed as part of standard clinical care.

plwALS were categorized by phenotype as classic ALS, bulbar ALS, respiratory ALS, flail arm, flail leg, and predominant upper motor neuron ALS^26^. Disease progression rate (PR) and progression rate without respiratory items (PR_noresp) at T0 and at T6, and variation of MRC between T0 and T6 (ΔMRC) were calculated using standard formulae (details in **Supplementary methods**).

STROBE checklist for observational studies has been filled (see **Supplementary materials_STROBE checklist**).

### 2.3 Salivary melatonin sampling and assay

Participants maintained a regular sleep schedule for 14 days before testing and completed 2-week sleep diaries to determine habitual sleep onset (HSO). Six saliva samples were self-collected at home under controlled dim-light conditions (<10 lux) at predefined time points relative to HSO (−3, −2, −1, 0, +1, and +8 h) using Salivette® (Sarstedt) devices. Samples were refrigerated after collection, processed, and stored at −80°C until analysis. Salivary MLT concentrations were measured by competitive ELISA (DRG SLV-4779). Additional sampling and assay details are provided in **Supplementary methods**.

### 2.4 MLT curve shaping

MLT phase and curve-shape metrics were derived using a reproducible R-based algorithm. Individual salivary concentration-time series, referenced to HSO, were modelled using cubic smoothing splines. DLMO was defined as the earliest time on the ascending limb at which the fitted curve reached 3 pg/mL.

DLMO_sleep_onset (time between the DLMO and the HSO time) and exploratory curve-shape metrics in the 3 hours after DLMO (MLT_DLMO2, MLT_DLMO3, AUC2, AUC3, slope) were also calculated. Only profiles with at least four non-missing samples and exceeding the 3 pg/mL threshold were retained.

Full computational details are reported in **Supplementary methods**.

Graphical representations of MLT secretion curves and their derived parameters at T0 and at T6, respectively, in **Figure 1S** and **Figure 2S** in **Supplementary materials**.

**Figure 1.**
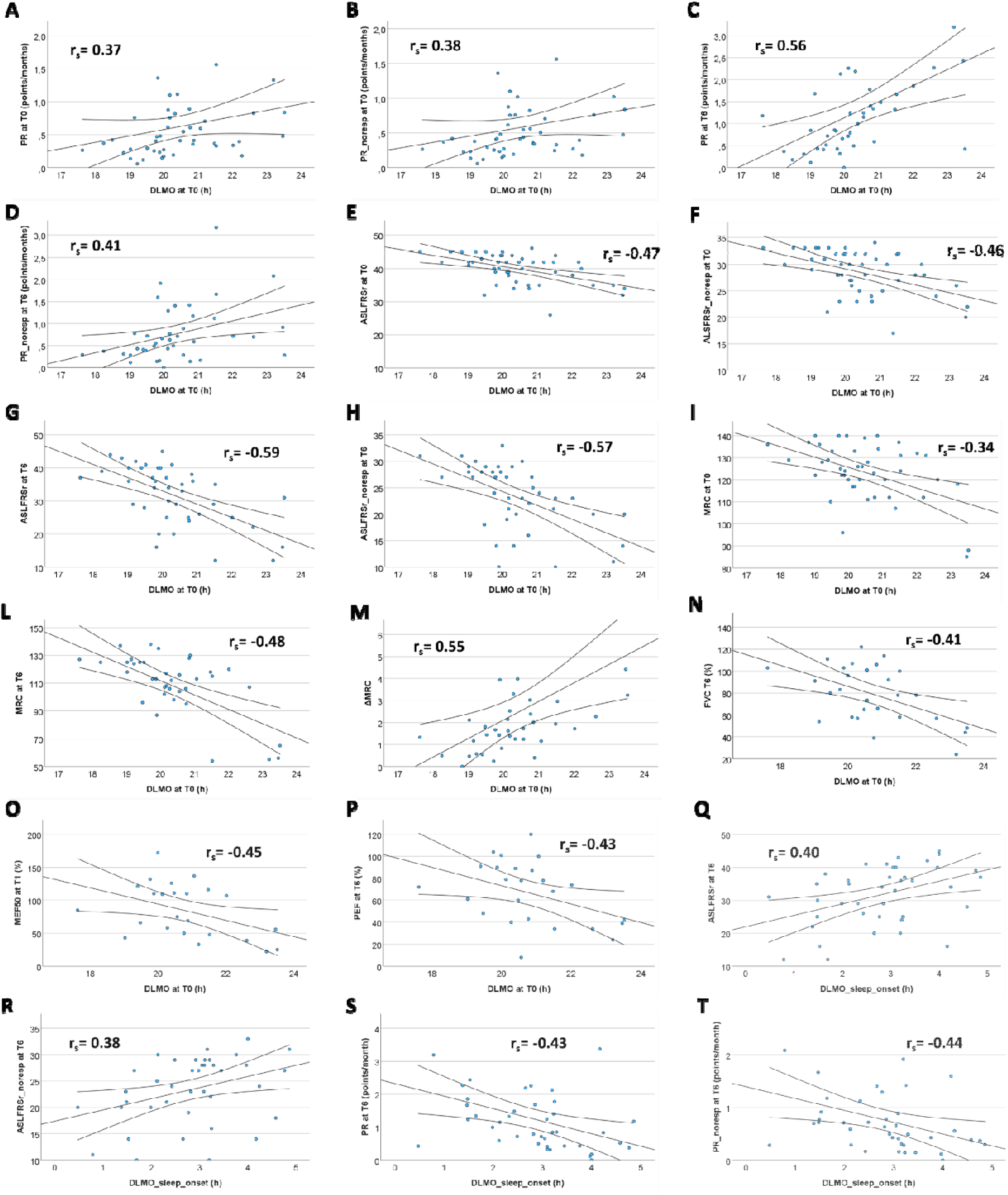
Correlation between clinical parameters and both DLMO and DLMO_sleep_onset in plwALS. <u>Legend</u>: DLMO: dim-light MLT onset; DLMO_sleep_onset: time between DLMO and sleep onset; ALSFRSr at T0: ALS Functional Rating Scale-revised total score at baseline; ALSFRSr_noresp at T0: ALSFRSr without the respiratory item at baseline; ALSFRSr at T6: ALS Functional Rating Scale-revised total score at month 6; ALSFRSr_noresp at T6: ALSFRSr without the respiratory item at month 6; MRC: Medical Research Council scale; ΔMRC: variation of MRC between T0 and T6; FVC: Forced Vital Capacity; MEF50: Maximum Expiratory Flow at 50%; PEF: Peak Expiratory Flow; T0: at the moment of sampling; T6: after 6 months from sampling.

**Figure 2.**
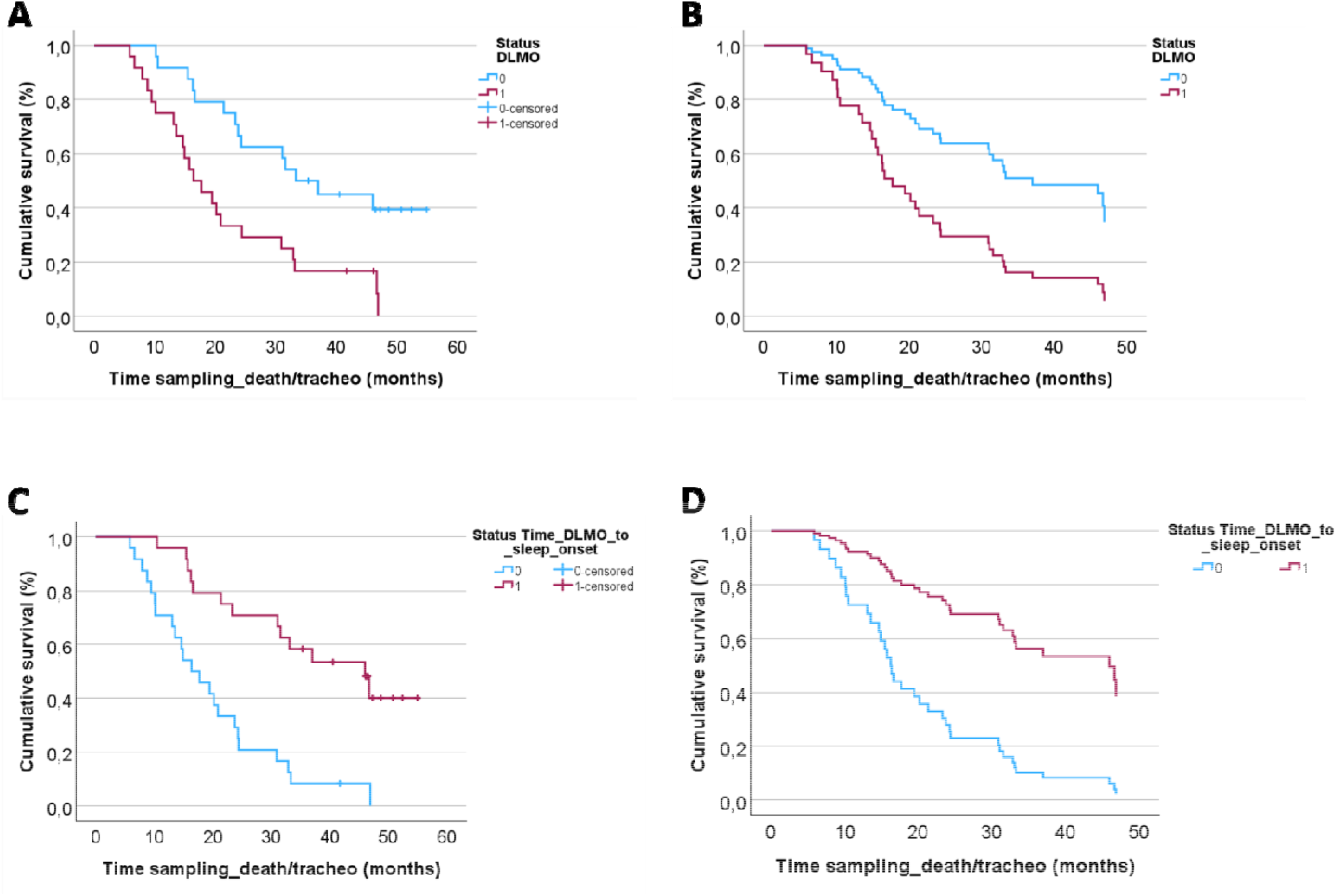
Survival analysis. (**A**) and (**C**) represent the Kaplan-Meier analysis, respectively categorizing by median DLMO and DLMO_sleep_onset. (**B**) shows survival curves covarying for sex, age at blood sampling, site of onset of the disease, and KINGS, categorizing DLMO using median (**D**) shows survival curves covarying for DLMO_sleep_onset, sex, age at blood sampling, site of onset of the disease, and KINGS categorizing DLMO_sleep_onset using median. <u>Legend</u>: in (A) and (B) light-blue line (status DLMO 0) correspond to time of DLMO ≤20:11, while red line (status DLMO 1) to time of DLMO >20:11; in (C) and (D) light-blue line (status DLMO_sleep_onset 0) correspond to levels of DLMO_sleep_onset ≤2:50 h, while red line (status DLMO_sleep_onset 1) to levels of DLMO_sleep_onset >2:50 h. Patients with later times of DLMO show a shorter survival. Patients with lower times of DLMO_sleep_onset show a shorter survival.

### 2.5 Statistical data analysis

Continuous variables are reported as mean ± standard deviation (SD); categorical variables as counts and percentages. Normality was assessed with Shapiro-Wilk tests: DLMO and DLMO_sleep_onset were normally distributed, whereas AUC2, AUC3, slope, MLT_DLMO2, and MLT_DLMO3 were not. Between-group comparisons used independent-samples t tests or Mann-Whitney U tests, as appropriate. Correlations between MLT secretion-curve parameters and clinical, laboratory, and instrumental variables were evaluated using Pearson’s correlation coefficient (r) or Spearman’s rank correlation coefficient (r_s_) for continuous variables and using logistic regression for dichotomous variables. Multiple regression analyses were conducted.

Survival was defined as time from baseline sampling to death or tracheostomy, with censoring in February 2026. Kaplan–Meier survival curves were generated, and multivariable Cox proportional hazards regression was performed. We explored potential non-linear associations applying restricted cubic splines (3 knots). Longitudinal paired comparisons were assessed with the Wilcoxon signed-rank test. Statistical significance was set at two-sided p<0.05. All analyses were performed using SPSS Statistics, version 30 (IBM Corp., Armonk, NY, USA).

## 3. Results

Fifty plwALS (mean age 64.9±8.9 years; 30 men, 20 women) were enrolled. Fourteen (28%) showed a bulbar onset and 7 carried mutations in the four major ALS genes (5 C9orf72, 2 SOD1). Two plwALS were excluded from the analysis because their MLT levels never reached the 3 pg/mL threshold, precluding calculation of DLMO and DLMO-derived measures.

The 15 age- and sex-matched HCs, with a mean age of 64.7±9.7 years, were useful to validate MLT assay performance. Demographic and clinical characteristics are summarized in **Table 1**. Up to T6, no patients required NIV or percutaneous gastrostomy.

**Table 1.** Participant demographic and clinical information.

|  | **ALS**<br>**(48)** | **HCs tot**<br>**(48)** | **HCs MLT**<br>**(15)** | **p-value**<br>**(ALS vs HCs tot)** | **p-value**<br>**(ALS vs HCs MLT)** |
| --- | --- | --- | --- | --- | --- |
| **Sex (M/F)** | 30/18 | 29/19 | 8/7 | 0.83<sup>ξ</sup> | 0.53<sup>ξ</sup> |
| **Age at T0 ± SD (y)** | 64.9 ± 8.9 | 65.8±10.6 | 64.7 ± 9.7 | 0.65 | 0.94 |
| **Site of onset (Spinal / Bulbar)** | 34/14 | - | - |  |  |
| **Presence of genetic mutation** | 7 | - | - |  |  |
| **ALSFRSr_tot at T0 ± SD (points)** | 40.0 ± 4.4 | - | - |  |  |
| **PR at T0 ± SD (points lost/month)** | 0.62 ± 0.55 | - | - |  |  |
| **KINGS stage at T0 (0/1/2/3/4)** | 0/11/20/17/0 | - | - |  |  |
| **MiToS stage at T0 (0/1/2/3/4)** | 43/5/0/0/0 | - | - |  |  |
| **FVC at T0 ± SD (%)** | 93.5 ± 18.4 | - | - |  |  |
| **Chronotype (morning/intermediate/evening)** | 27/16/5 | 25/17/6 | 8/5/2 | 0.91<sup>ξ</sup> | 0.95<sup>ξ</sup> |
| **Habitual sleep onset time (h)** | 23:06 ± 0:43 | 23:10 ± 0:56 | 23:11 ± 1:04 | 0.70 | 0.79 |

<sup>ξ</sup> $\chi^2$ test.
Legend: ALSFRSr\_tot: ALS Functional Rating Scale-revised total score; PR: progression rate; FVC: Forced Vital Capacity; SD: Standard Deviation; n/a: not assessed.

### 3.1 Evaluation of salivary MLT in plwALS

Neither plwALS nor HCs showed significant day-to-day variation in HSO (p>0.1).

Among plwALS, 27/48 (56.3%) showed a morning chronotype, 5/48 (10.4%) an evening chronotype, and 16/48 (33.3%) an intermediate chronotype. No differences with HCs were found (p>0.1, **Table 1**).

Looking at the 15 HCs who provided saliva samples, their DLMO levels (20:58±0:50) were consistent with published values for the same age range^13,27^, supporting the overall assay plausibility.

Although this was not a diagnostic study, we observed a significant earlier DLMO in plwALS (20:24±1:18) compared both to our HCs (20:58±0:50; p=0.028) and also to literature data of DLMO^13^ in subjects aged 50 to 80 years (21:12±0:26; p=0.0002). No differences in HSO (p=0.79) and in chronotype (p=0.95) between plwALS and HCs.

No significant baseline DLMO differences were observed by sex (p=0.68), bulbar versus spinal onset (p=0.31), and genetic versus sporadic forms (p=0.26). However, inter-individual variability was greater in plwALS than in HCs, suggesting heterogeneous circadian-phase behaviour across patients.

### 3.2 Sleep Macrostructure in plwALS

Compared with the 48 age- and sex-matched HCs who underwent PSG, plwALS had similar total sleep time (367±64 vs 385±86; p=0.18), but lower sleep efficiency (79.5±9.2 vs 84.2±8.5; p=0.021), higher Deep Sleep percentage (27.1±11.7 vs 20.0±8.2; p=0.006), and lower REM sleep percentage (14.2±4.9 vs 19.1±4.4; p=0.005).

### 3.3 Correlation of salivary MLT with clinical, laboratory, and instrumental parameters in plwALS Later baseline DLMO correlated with greater baseline disease severity and faster progression

Specifically, DLMO at T0 correlated positively with PR at T0 (r_s_=0.37, p=0.01), PR_noresp at T0 (r_s_=0.38, p=0.009), PR at T6 (r_s_=0.56, p=1.4×10^−4^), and PR_noresp at T6 (r_s_=0.41, p=0.01) (**Figure 1A,B,C,D**). Moreover, DLMO at T0 correlated inversely with ALSFRSr at T0 (r_s_=-0.47, p=9.28×10^−4^; **Figure 1E**), with ALSFRSr_noresp at T0 (r_s_=-0.46, p=0.001; **Figure 1F**), with ALSFRSr at T6 (r_s_=-0.59, p=4.77×10^−5^; **Figure 1G**), and with ALSFRSr_noresp at T6 (r_s_=-0.57, p=1.17×10^−4^; **Figure 1H**). DLMO also correlated inversely with MRC at T0 and at T6 (respectively, r_s_=-0.34, p=0.021 and r_s_=-0.48, p=0.002; **Figure 1I,L**), while directly with ΔMRC (r_s_=0.55, p=3.64×10^−4^; **Figure 1M**). All relationships remained significant after adjustment for sex, age, and site of onset (**Supplementary materials, Table 1S**).

Respiratory outcomes showed a similar pattern. By 6 months, 32 plwALS had developed dyspnoea and/or orthopnoea. A later baseline DLMO independently predicted this outcome (OR 2.46, 95%CI 1.021-5.00; p=0.013); statistically significance maintained also after multiple adjustments (**Supplementary materials, Table 3S**). Consistently, DLMO at T0 correlated inversely with FVC at T6 (r=−0.41; p=0.031; **Figure 1N**), MEF50 at T6 (r=−0.45; p=0.027; **Figure 1O**), and PEF at T6 (r =−0.43; p=0.033; **Figure 1P**); significance confirmed these correlations remained significant after adjustment for sex, age, ALSFRSr at T0, and site of onset (**Supplementary materials, Table 1S**).

No association (p>0.1) of DLMO and other MLT curves-derived parameters with PSG and ABG at T0 and at T6 were found. Likewise, no other relevant associations were observed for AUC2, AUC3, slope, MLT_DLMO2, or MLT_DLMO3.

For DLMO_sleep_onset, a positive direct correlation was found both with ALSFRSr at T6 (r=0.40, p=0.01; **Figure 1Q**) and with ALSFRSr_noresp at T6 (r=0.38, p=0.017; **Figure 1R**). Moreover, DLMO_sleep_onset at T0 correlated inversely with PR at T6 (rs=-0.43, p=0.005) and PR_noresp at T6 (r=-0.44, p=0.004) (**Figure 1S,T**). All relationships remained significant after adjustment for sex, age, and site of onset (**Supplementary materials, Table 2S**).

### 3.4 Survival analysis

Among the 48 plwALS included in the analysis, 36 had either deceased or undergone tracheostomy by February 2026. Interim survival analyses were therefore performed, acknowledging the limited sample size. Given the observed correlation of DLMO and of DLMO_sleep_onset with motor and functional disease progression, Kaplan-Meier curves were generated. DLMO was dichotomized at the median (≤20:11 time vs >20:11 time). Survival diverged significantly in favour of earlier DLMO (Log-Rank test, χ^2^=9.707, p=0.002; Breslow test, χ^2^=9.230, p=0.002; Tarone-Ware test, χ^2^=9.407, p=0.002, **Figure 2A**), and this remained significant after stratification by site of onset (Log-Rank test, χ^2^=8.780, p=0.003; Breslow test, χ^2^=8.225, p=0.004; Tarone-Ware test, χ^2^=8.432, p=0.004).

Univariate and multivariate Cox regression analyses were conducted for both continuous and categorical DLMO at T0 (**Supplementary materials, Table 4S**). Later DLMO was associated with shorter survival, both as a continuous (χ^2^=27.503, p=1.07×10^−6^) and categorical (χ^2^= 9.255, p=0.002) variable, after correction for age, sex, site of onset, and disease stage. In a sensitivity analysis including chronotype, the association remained significant. Multivariable Cox-derived curves are shown in **Figure 2B**.

Looking at DLMO_sleep_onset values, they were dichotomized at the median (≤2:50 h vs. >2:50 h). Lower values, indicating a shorter interval between DLMO and sleep onset, were associated with shorter survival (Log-Rank test, χ^2^=16.854, p=4.04×10^−5^; Breslow test, χ^2^=15.021, p=0.0001; Tarone-Ware test, χ^2^=16.030, p=6.23×10^−5^, **Figure 2C**). This significance persisted when further stratified by site of onset (Log-Rank test, χ^2^=12.932, p=0.0003; Breslow test, χ^2^=11.545, p=0.0003; Tarone-Ware test, χ^2^=12.559, p=0.0004). Univariate and multivariate Cox regression analyses were conducted for both continuous and categorical DLMO_sleep_onset at T0 (**Supplementary materials, Table 5S**). Shorter DLMO_sleep_onset remained associated with lower survival both as a continuous (χ^2^=13.396 p=2.52×10^−4^) and categorical (χ^2^= 16.435, p=5.04×10^−5^) variable after correction for age, sex, site of onset, and disease stage. Multivariable Cox-derived curves are shown in **Figure 2D**.

Restricted cubic spline analyses further supported these findings for both DLMO and DLMO_sleep_onset, with no evidence of non-linearity (DLMO: overall p=1.35×10^−5^, non-linearity p=0.991; DLMO_sleep_onset: overall p=9.83×10^−4^, non-linearity p=0.854; **Supplementary materials, Table 6S**).

### 3.5 Longitudinal evaluation

No baseline differences in clinical and demographic data were observed (Mann-Whitney p>0.1, **Table 2**) between the full cohort of 48 plwALS and the subgroup of 28 with repeat DLMO assessment at approximately 6 months (mean 6.63±0.78; median 6.55 [IQR 6.12–6.77]). Major clinical parameters at T0 and T6 for these 28 plwALS are shown in **Supplementary materials, Table 7S**. Longitudinal analysis showed no significant changes in DLMO, DLMO_sleep_onset, and other curve-shape metrics between T0 and T6 (p>0.1).

**Table 2.**
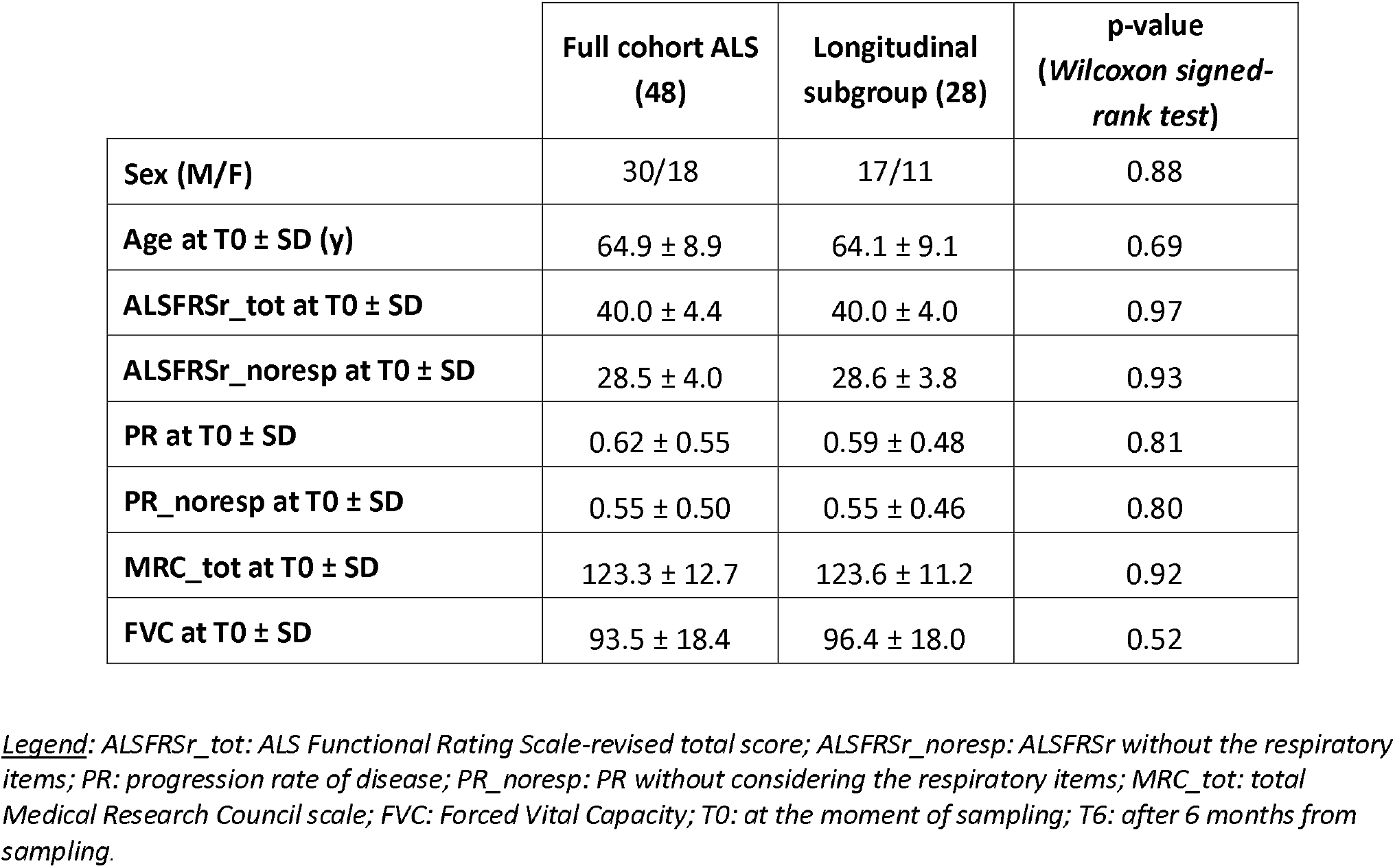
Baseline clinical parameters and scales in full cohort of 48 plwALS and in the subgroup of 28 who underwent longitudinal sampling.

## Discussion

In this prospective cohort of early-stage plwALS, we found circadian phase abnormalities that carry clinically relevant information. First, DLMO timing in plwALS differed from matched HCs and from large published normative datasets^13^, indicating that MLT rhythm is perturbed early in ALS rather than emerging only as a late consequence of respiratory failure. Second, within the ALS cohort, later DLMO and shorter DLMO_sleep_onset interval were associated with worse functional status, faster short-term progression, incident respiratory symptoms, and poorer survival. Third, phase-based measures (DLMO and DLMO_sleep_onset) outperformed exploratory curve-shape metrics, suggesting that circadian timing, rather than early-night MLT amplitude, areas or slope, carries the most clinically relevant signal in this dataset.

Circadian disruption and altered MLT physiology recur across neurodegenerative diseases, supporting the plausibility that ALS, includes early network vulnerabilities affecting circadian control^15,16,28^. To our knowledge, this is the first study to objectively characterize endogenous salivary MLT timing in ALS, and to demonstrate longitudinal associations with motor and functional decline.

The shifted DLMO observed in early ALS, together with greater inter-individual variability, is consistent with early circadian network vulnerability and heterogeneous compensatory responses. These findings should be interpreted alongside our observation of increased slow-wave sleep, reduced REM sleep, and lower sleep efficiency in the ALS cohort. Sleep macrostructure changes may reflect a less restorative sleep in plwALS; accordingly, the phase-advanced DLMO in early-ALS could represent a compensatory shift in sleep–wake regulatory circuits in response to increased sleep pressure. This hypothesis is supported by studies in which both pharmacological^29^ and environmental^30^ operations ameliorate sleep quality and induce an earlier DLMO.

A further relevant finding is the association between delayed DLMO and subsequent respiratory symptom development despite weak baseline correlations with spirometry or PSG metrics. This suggests that MLT timing may capture a more upstream and integrative dimension of network dysfunction than conventional respiratory measures. Whereas spirometry is relatively late and effort-dependent and PSG provides a single-night snapshot, DLMO reflects central circadian phase and may integrate cumulative influences such as light exposure, inflammation, metabolic stress, and hypoxic burden, potentially anticipating overt ventilatory decline. Conversely, circadian misalignment may itself contribute to impaired ventilatory control and symptom emergence^31^. This interpretation is supported by evidence from non-neuromuscular respiratory disorders, including OSA, in which circadian disruption and clock-gene abnormalities may partially improve with treatment^32,33^. Our findings extend this framework to ALS by identifying endogenous MLT phase as a correlate of respiratory symptom trajectories and survival.^28^.

Beyond chronobiology, MLT also has antioxidant, anti-inflammatory, mitochondrial, and proteostatic actions relevant to motor neuron vulnerability. In preclinical ALS models, MLT engages SIRT1 signaling and autophagy, reduces apoptotic cascades, and may counter secondary injury pathways, positioning MLT system at the intersection of timing and neuroprotection^34^. While our study was not designed to test treatment effects, the combination of altered phase and plausible neuronal actions motivates future trials that are explicitly timed relative to endogenous phase.

### Biological plausibility

Converging evidence indicates that non-motor symptoms, including sleep and circadian complaints, are clinically relevant in ALS even early in the disease course^3^. In parallel, recent imaging studies link hypothalamic atrophy to weight loss, behavioral features, and survival, indicating that the hypothalamus is not merely epiphenomenally involved^35^. Because the suprachiasmatic nucleus (SCN) lies within the anterior hypothalamus and circadian outputs rely on hypothalamic integrity, circadian phase abnormalities in ALS are biologically plausible^11^. Moreover, glymphatic-lymphatic waste clearance is under circadian control and peaks during the biological night; mistimed sleep could impair clearance and exacerbate neuroinflammatory processes to which MLT is mechanistically linked^36^.

The association between delayed DLMO and worse outcomes may reflect several interacting mechanisms. Central circadian dysregulation related to hypothalamic neuronal loss and TDP-43 pathology, including orexinergic and other neuropeptidergic systems, may desynchronize SCN signaling and mis-time pineal MLT output. Circadian misalignment may also impair sleep-dependent restorative and clearance functions, promoting toxic metabolite accumulation and neuroinflammation. Respiratory factors may further contribute bidirectionally: intermittent hypoxaemia and sleep fragmentation may shift circadian phase, while circadian disorganisation may further destabilise respiratory control. These mechanisms are not mutually exclusive and likely reinforce one another across disease progression.

### Translational and Therapeutic implications

If replicated in larger multi-center cohorts, DLMO and DLMO_sleep_onset could support risk stratification and trial enrichment in early-ALS, complementing established clinical staging and emerging fluid biomarkers^1^. Because circadian interventions are feasible and scalable, including timed light exposure, activity scheduling, and appropriately timed low-dose MLT in selected circadian disorders, our findings support future ALS studies incorporating objective circadian phenotyping and testing whether improving circadian alignment favourably affects symptoms or downstream physiology^28,29^. Moreover, since NIV improves survival^37,38^, DLMO might help identify patients who could benefit from earlier NIV initiation, pending validation in larger cohorts.

The broader MLT literature also warrants careful interpretation. Preclinical ALS models suggest MLT can modulate pathways relevant to motor neuron vulnerability, including apoptosis and autophagy^18,19^. Observational human data, including registry^20^ and clinical-trial databases^34^, have suggested associations between MLT use and slower progression, but such findings require confirmation in prospective, multicenter, phase-anchored clinical trials.

### Strengths, limitations, and future directions

Limitations include modest sample size (especially for HCs and the 6-month re-sampling subset), monocenter design of the study, potential residual confounding, and the inherent variability of home-based light exposure and sampling behavior. The follow-up window may be too short to capture slower within-subject phase drift, seasonal effects, or later-stage circadian deterioration. Additionally, while DLMO is robust, broader characterization (e.g., 24-hour MLT profiles, actigraphy, objective light sensing, and repeated multi-night collections) would help separate reduced amplitude from phase shifts and clarify how circadian biology changes across disease stages. Moreover, another limitation is the absence of additional fluid biomarkers concurrently measured (such as NfL and pNfH).

Strengths include: (i) a prospective design linking baseline DLMO to prespecified clinical outcomes (ALSFRSr, progression rate, incident respiratory symptoms, and survival); (ii) rigorous, home-based saliva collection under dim light with validated thresholds; and (iii) concurrent respiratory and sleep phenotyping to situate DLMO within standard ALS care. These elements adhere to current best practices for circadian biomarker deployment in clinical research^12^.

Future work should: (1) replicate these findings in multi-center cohorts; (2) integrate objective sleep–circadian–respiratory monitoring (actigraphy, light sensors, PSG); (3) link DLMO to neuroimaging markers of hypothalamic and brainstem network integrity; and (4) asses interventional trials using personalized circadian strategies (timed light, activity, and MLT).

## Conclusions

Overall, our findings support the presence of circadian timing abnormalities from the early stages of ALS and suggest prognostic relevance. Later baseline DLMO was associated with faster disease progression, greater functional decline, incident respiratory symptoms, and poorer survival, independent of standard covariates and not explained by conventional respiratory or PSG measures in this cohort. Together, these data position DLMO as a promising noninvasive, physiology-anchored prognostic biomarker in early-ALS. If validated, DLMO could become a practical, noninvasive circadian biomarker to complement the ALS prognostic toolkit and guide future phase-targeted supportive trials.

## Data Availability

All data produced in the present study are available upon reasonable request to the authors

