## Supplementary methods for "“Salivary dim-light melatonin onset in early Amyotrophic Lateral Sclerosis predicts functional decline, respiratory symptom emergence, and survival”"

**2.2 Clinical assessment**

Disease progression rate (PR) and progression rate without respiratory items (PR_noresp) at T0 and at T6 were calculated as follows:

$$PR\_T0=\frac{48-\mathrm{ALSFRSr}_{T0}}{Date T0 -Date symptom onset}$$

$$PR\_noresp\_T0=\frac{36-{ALSFRSr\_noresp}_{T0}}{Date T0 -Date symptom onset}$$

$$PR\_T6=\frac{\mathrm{ALSFRSr}_{T0}-\mathrm{ALSFRSr}_{T6}}{Date T6 -Date T0}$$

$$PR\_noresp\_T6=\frac{{ALSFRSr\_noresp}_{T0}-{ALSFRSr\_noresp}_{T6}}{Date T6 -Date T0}$$

Variation of MRC between T0 and T6 (ΔMRC) was calculated as follows:

$$\Delta MRC=\frac{\mathrm{MRC}_{T0}-\mathrm{MRC}_{T6}}{Date T6 -Date T0}$$

**2.3 Salivary melatonin sampling and assay**

Participants were instructed to maintain a regular (±30 minutes) sleep schedule for 14 days prior to testing and to complete sleep diaries at home for two weeks, maintaining their usual living conditions, to determine habitual sleep onset time (HSO). On the sampling day, six saliva samples were self-collected at home under dim-light conditions (<10 lux), using Salivette® (Sarstedt AG & Co. KG, Germany): 3, 2, and 1 hour before HSO, at HSO, 1 hour after HSO, and 8 hours after HSO. Light exposure was monitored pragmatically by caregivers using a smartphone lux-meter application (Photone – Grow Light Meter; Lightray Innovation GmbH, Zurich, Switzerland), and participants completed a diary of naps, physical activity, and evening behaviours. Participants rinsed the mouth 15 minutes before sampling, placed the swab sublingually for 2-3 minutes and then in the supplied tube labelled with date, time and participant code. Each sample was refrigerated immediately after collection.

Samples were centrifuged at 2500 g for 10 minutes at 4 °C, aliquoted into cryovials, and stored at −80 °C until analysis was performed.

Before ELISA testing, samples were thawed at 4°C overnight, brought to room temperature for 30 minutes in a dark box, vortexed, and centrifuged at 1800 g for 10 minutes before analysis to deposit potential residual sediments.

MLT concentrations were determined by competitive ELISA (SLV-4779 Kit, DRG Instruments GmbH, Germany) according to manufacturer instructions. Optical density was read at 450 nm within 15 minutes. Plates were read using a CLARIOstar Plus plate reader (BMG LABTECH, Ortenberg, Germany), standard curves were fitted using 4-parameter logistic regression with MARS data analysis software (R²≥0.99), and sample MLT concentrations were interpolated accordingly. According to manufacturer’s instructions, analytical sensitivity was set <0.854 pg/mL. The median intra-assay and inter-assay coefficients of variation were below 15% for all the assays.

**2.4 MLT curve shaping**

A reproducible algorithm was implemented in R to derive MLT phase and shape metrics from each subject’s salivary series. Sampling times were referenced to HSO.

For each individual, the concentration–time profile was modelled with a cubic smoothing spline fitted to all non-missing points. The fitted function y^(t) was then sampled on fine grids (step 0.01 hour) to obtain the following quantities:

- **DLMO (h):** identified on the ascending limb as the earliest time at which y^(t) reached the MLT salivary threshold of 3 pg/mL.
- **MLT level at DLMO+2 (MLT_DLMO2) and at DLMO+3 (MLT_DLMO3) (pg/mL):** respectively levels of MLT at 2 and 3 hours after DLMO.
- **Mean slope from DLMO to DLMO+2 (slope) (pg/[mL*h]):** a finite-difference estimate of the average rise over the first 2 hours after DLMO;
- **Area under the curve between DLMO and DLMO+2 (AUC2) and DLMO+3 (AUC3) (pg*h/mL):** numerically integrated using a rectangular rule with Δt=0.01 hours, representing the MLT secretion during the first 2 and first 3 hours after DLMO, respectively.

We also calculated the **time between DLMO and sleep onset (DLMO_sleep_onset) (h)**, defined as the elapsed time between the DLMO and the HSO time.

Only profiles with at least four non-missing MLT samples and which exceeded the threshold of 3 pg/mL were retained for analysis.

All graphical representations of MLT secretion curves and their derived parameters at T0 and at T6 are, respectively, in **Figure 1S** and in **Figure 2S** in **Supplementary material**.
