## Supplementary materials for "“Salivary dim-light melatonin onset in early Amyotrophic Lateral Sclerosis predicts functional decline, respiratory symptom emergence, and survival”"

**Supplementary data**

**Table 1S - Multiple regression analysis between DLMO and clinical parameters**

| **Dependent variable ^δ^** | **Adjustments*** | **R of the model** | **F** | **p-value** |
| --- | --- | --- | --- | --- |
| ***ALSFRSr at T0*** | Age at saliva sampling (cont), sex (M/F), site of onset (S/B) | -0.490 | 14.25 | **4.66x 10^-4^** |
| ***ALSFRSr_noresp at T0*** |  | -0.477 | 13.24 | **7.04x 10^-4^** |
| ***MRC at T0*** |  | -0.647 | 15.14 | **1.12x 10^-5^** |
| ***ALSFRSr at T6*** |  | -0.594 | 21.24 | **4.27x 10^-5^** |
| ***ALSFRSr_noresp at T6*** |  | -0.549 | 16.36 | **2.47x 10^-4^** |
| ***PR at T6*** |  | 0.533 | 7.54 | **0.002** |
| ***PR_noresp at T6*** |  | 0.364 | 5.82 | **0.02** |
| ***MRC at T6*** |  | -0.639 | 12.09 | **1.02x 10^-4^** |
| ***ΔMRC at T0-T6*** |  | 0.436 | 8.45 | **0.006** |
| ***FVC at T6*** | Age at blood sampling (cont), sex (M/F), site of onset (S/B, and ALSFRSr at T0 | -0.512 | 9.25 | **0.005** |
| ***MEF50 at T6*** |  | -0.457 | 5.81 | **0.025** |
| ***PEF at T6*** |  | -0.439 | 5.49 | **0.028** |

*Method used for inserting variables in the model: Stepwise Backwards

*Legend: ALSFRSr: ALS Functional Rating Scale-revised total score; ALSFRSr_noresp: ALSFRSr without the respiratory items; PR: progression rate of disease;* *PR_noresp: PR without considering the respiratory items; ΔMRC: change of* *Medical Research Council scale between T0 and T6; FVC: Forced Vital Capacity; MEF50: Maximum Expiratory Flow at 50%; PEF: Peak Expiratory Flow; T0: at the moment of sampling; T6: after 6 months from sampling.*

**Table 2S - Multiple regression analysis between DLMO_sleep_onset and clinical parameters**

| **Dependent variable ^δ^** | **Adjustments*** | **R of the model** | **F** | **p-value** |
| --- | --- | --- | --- | --- |
| ***ALSFRSr at T6*** | Age at saliva sampling (cont), sex (M/F), site of onset (S/B) | 0.412 | 7.96 | **0.007** |
| ***ALSFRSr_noresp at T6*** |  | 0.362 | 5.72 | **0.022** |
| ***PR at T6*** |  | -0.397 | 7.31 | **0.010** |
| ***PR_noresp at T6*** |  | -0.367 | 5.92 | **0.02** |

*Method used for inserting variables in the model: Stepwise Backwards

*Legend: ALSFRSr: ALS Functional Rating Scale-revised total score; ALSFRSr_noresp: ALSFRSr without the respiratory items; PR: progression rate of disease;* *PR_noresp: PR without considering the respiratory items; T0: at the moment of sampling; T6: after 6 months from sampling.*

**Table 3S - Binary logistic regression models testing whether baseline DLMO predicts incident respiratory symptoms at 6 months**

| **Model** | **OR for DLMO at T0 (95% CI)** | **p value** |
| --- | --- | --- |
| **Univariable** | 2.46 (1.21-5-00) | **0.013** |
| **Adjusted for age, sex, and site of onset** | 3.24 (1.38-7.60) | **0.006** |
| **Adjusted for age, sex, site of onset, and FVC at T0** | 3.20 (1.35-7.61) | **0.008** |
| **Adjusted for age, sex, site of onset, and KINGS at T0 (>2 vs ≤2)** | 2.88 (1.23-6.72) | **0.015** |
| **Adjusted for age, sex, site of onset, KINGS at T0 (>2 vs ≤2), and FVC at T0** | 3.11 (1.21-8.04) | **0.019** |

Legend: DLMO was analysed as a continuous variable; odds ratios (OR) refer to each 1-hour delay in baseline DLMO.

Site of onset was entered as bulbar versus spinal. CI: confidence interval; FVC: forced vital capacity.

**Table 4S - Cox proportional hazard models using DLMO**

|  | **Univariate Analysis— Survival after sampling** | | |
| --- | --- | --- | --- |
|  |  | **HR (95% CI)** | **p** |
|  | **Age at blood sampling (continuous)** | 1.029 (0.994-1.066) | 0.105 |
|  | **Sex (M vs F)** | 1.221 (0.627-2.375) | 0.557 |
|  | **Site of onset (Spinal/Bulbar)** | 1.641 (0.777-3.465) | 0.190 |
|  | **KINGS (categorial – ≤2 vs >2)** | 1.670 (0.846-3.294) | 0.148 |
|  | **Morningness-Eveningness (continuous)** | 0.974 (0.891-1.065) | 0.567 |
|  | **DLMO (categorial - upper/lower median)** | 2.783 (1.316-5.882) | **0.007** |
|  | **DLMO (continuous)** | 1.945 (1.408-2.685) | **5.32 x10^-5** |
|  | **Multivariate analysis*—Survival after sampling**  *Adjusting for DLMO, age (cont), sex (M/F), site of onset (S/B), and KINGS* | | |
| **Statistic of the model - DLMO (categorial - median)** | | | **0.002** |
| **DLMO (median)** | | 2.778 (1.400-5.511) | **0.003** |
| **Statistic of the model - DLMO (continuous)** | | | **1.07 x10^-6^** |
| **DLMO (continuous)** | | 2.107 (1.548-2.866) | **1.07 x10^-6^** |
|  | **Multivariate analysis* (sensitivity analysis) —Survival after sampling**  *Adjusting for DLMO, age (cont), sex (M/F), site of onset (S/B), and*  *Morningness-Eveningness (cont)* | | |
| **Statistic of the model - DLMO (categorial - median)** | | | **0.004** |
| **DLMO (median)** | | 2.708 (1.341-5.468) | **0.005** |
| **Statistic of the model - DLMO (continuous)** | | | **1.53x10^-6^** |
| **DLMO (continuous)** | | 2.193 (1.579-3.045) | **2.11 x10^-6^** |

*Method used for inserting variables in the model: Stepwise Backwards Regression

**Table 5S - Cox proportional hazard models using DLMO_sleep_onset**

|  | **Univariate Analysis—Survival after sampling** | | |
| --- | --- | --- | --- |
|  |  | **HR (95% CI)** | **p** |
|  | **Age at blood sampling (continuous)** | 1.029 (0.994-1.066) | 0.105 |
|  | **Sex** | 1.221 (0.627-2.375) | 0.557 |
|  | **Site of onset (Spinal/Bulbar)** | 1.641 (0.777-3.465) | 0.190 |
|  | **KINGS (categorial – ≤2 vs >2)** | 1.670 (0.846-3.294) | 0.148 |
|  | **DLMO_sleep_onset (categorial - upper/lower median)** | 0.249 (0.123-0.506) | **4.04 x10^-5^** |
|  | **DLMO_sleep_onset (continuous)** | 0.537 (0.385-0.750) | **2.18 x10^-4^** |
|  | **Multivariate analysis*—Survival after sampling**  *Adjusting for DLMO_sleep_onset, age (cont), sex (M/F), site of onset (S/B), and KINGS* | | |
| **Statistic of the model - DLMO_sleep_onset (categorial - median)** | | | **5.04 x10^-5^** |
| **DLMO_sleep_onset (median)** | | 0.252 (0.124-0.513) | **1.45 x10^-4^** |
| **Statistic of the model - DLMO_sleep_onset (continuous)** | | | **2.52 x10^-4^** |
| **DLMO_sleep_onset (continuous)** | | 0.531 (0.377-0.748) | **2.87x10^-4^** |

*Method used for inserting variables in the model: Stepwise Backwards Regression.

**Table 6S - Restricted cubic spline Cox models with 3 knots**

| **Analysis** | **Adjustments** | **Knots (hours)** | **Overall association p-value** | **Non-linearity p-value** |
| --- | --- | --- | --- | --- |
| **DLMO continuous (main analysis)** | Age (continuous), sex (M/F), site of onset (spinal/bulbar), KINGS at T0 (≤2 vs >2) | 19.03;  20.23;  22.22 | **1.35 × 10^-5** | 0.991 |
| **DLMO continuous (sensitivity analysis)** | Age (continuous), sex (M/F), site of onset (spinal/bulbar), KINGS at T0 (≤2 vs >2), chronotype (continuous) | 19.08;  20.28;  22.25 | **2.99 × 10^-5** | 0.679 |
| **DLMO_sleep_onset continuous**  **(main analysis)** | Age (continuous), sex (M/F), site of onset (spinal/bulbar), KINGS at T0 (≤2 vs >2) | 1.44;  2.84;  4.05 | **9.83 × 10^-4** | 0.854 |

Cox proportional hazards models were fitted using restricted cubic splines with 3 knots to explore potential non-linear associations between baseline DLMO, baseline DLMO_sleep_onset, and survival after sampling. Models were adjusted for age, sex, site of onset, and baseline KINGS stage (≤2 vs >2); for DLMO, an additional sensitivity analysis further adjusted for chronotype. The overall association p-value tests whether the predictor is associated with survival, whereas the non-linearity p-value tests whether the association deviates significantly from linearity.

**Table 7S – Clinical parameters and scales in ALS patients who underwent longitudinal sampling**

|  | **ALS (28)** | |  |
| --- | --- | --- | --- |
|  | **T0** | **T6** | **p-value**  **(*Wilcoxon signed-rank test*)** |
| **ALSFRSr_tot ± SD** | 40.0 ± 4.4 | 31.8 ± 8.9 | **3.48 x 10^-8^** |
| **ALSFRSr_noresp ± SD** | 28.5 ± 4.0 | 23.3 ± 6.2 | **5.01 x 10^-8^** |
| **PR ± SD** | 0.61 ± 0.55 | 1.24 ± 1.02 | **1.09 x 10^-5^** |
| **PR_noresp ± SD** | 0.57 ± 0.50 | 0.77 ± 0.67 | **0.016** |
| **MRC_tot ± SD** | 123.3 ± 12.9 | 107.8 ± 23.8 | **1.12 x 10^-7^** |
| **FVC ± SD** | 93.5 ± 18.6 | 78.8 ± 25.8 | **1.15 x 10^-5^** |

*Legend: ALSFRSr_tot: ALS Functional Rating Scale-revised total score; ALSFRSr_noresp: ALSFRSr without the respiratory items; PR: progression rate of disease; PR_noresp: PR without considering the respiratory items; MRC_tot: total Medical Research Council scale; FVC: Forced Vital Capacity; T0: at the moment of sampling; T6: after 6 months from sampling.*

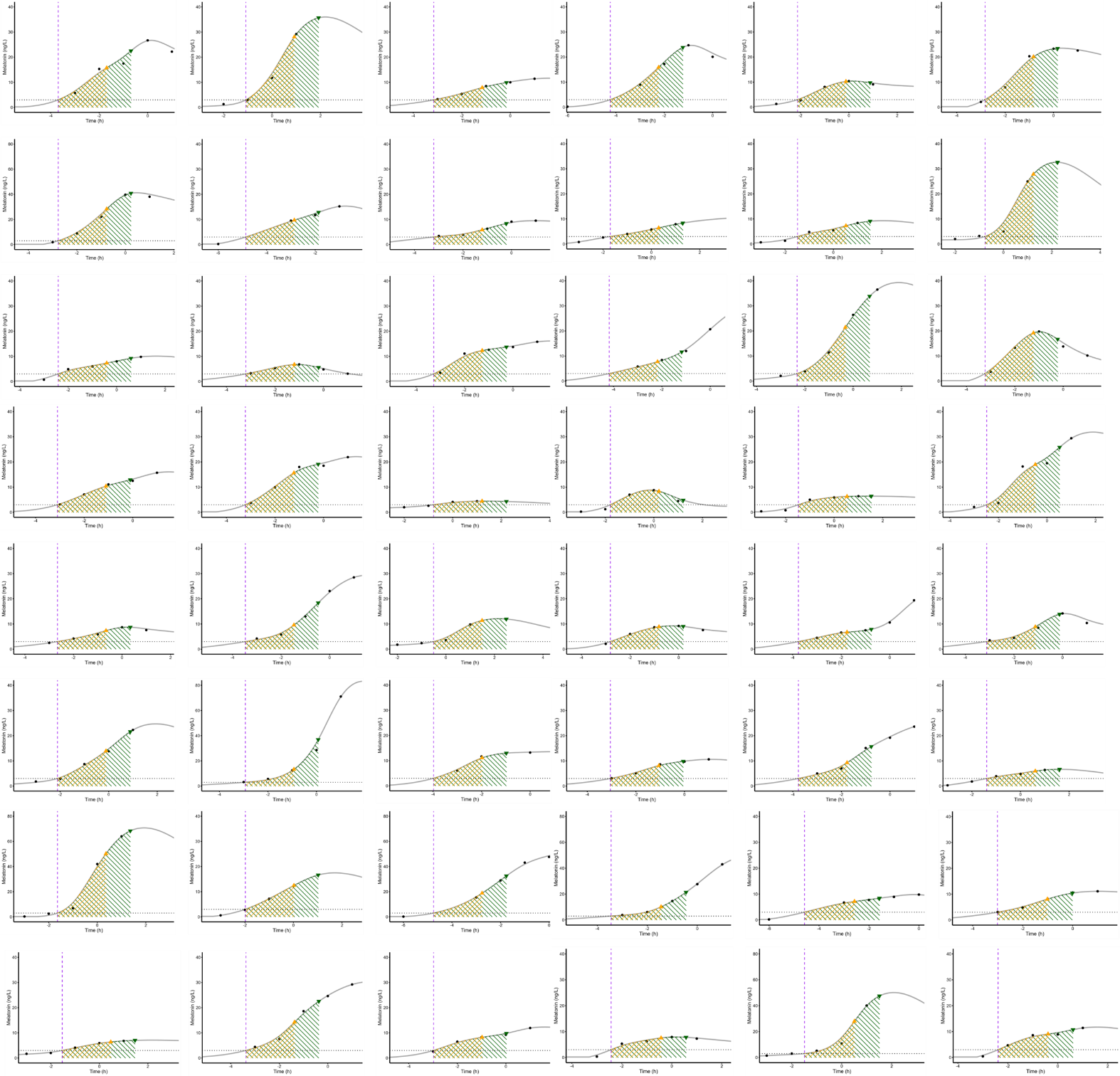

**Figure 1S – MLT secretion curves of plwALS at T0 and derived parameters**

*Legend: purple vertical dash line = DLMO; grey horizontal dot line = MLT levels of 3 pg/ml; black dots = levels of MLT at different time points (at 3, 2, and 1 hours before habitual sleep onset [HSO], at HSO, 1 hour after, and 8 hours after HSO); grey line = line of interpolation of different MLT points; orange triangle = MLT level at DLMO+2 hours (MLT_DLMO2); green triangle = MLT level at DLMO+3 hours (MLT_DLMO3); hatched orange area = Area under the curve between DLMO and DLMO+2; hatched green area = Area under the curve between DLMO and DLMO+3.*

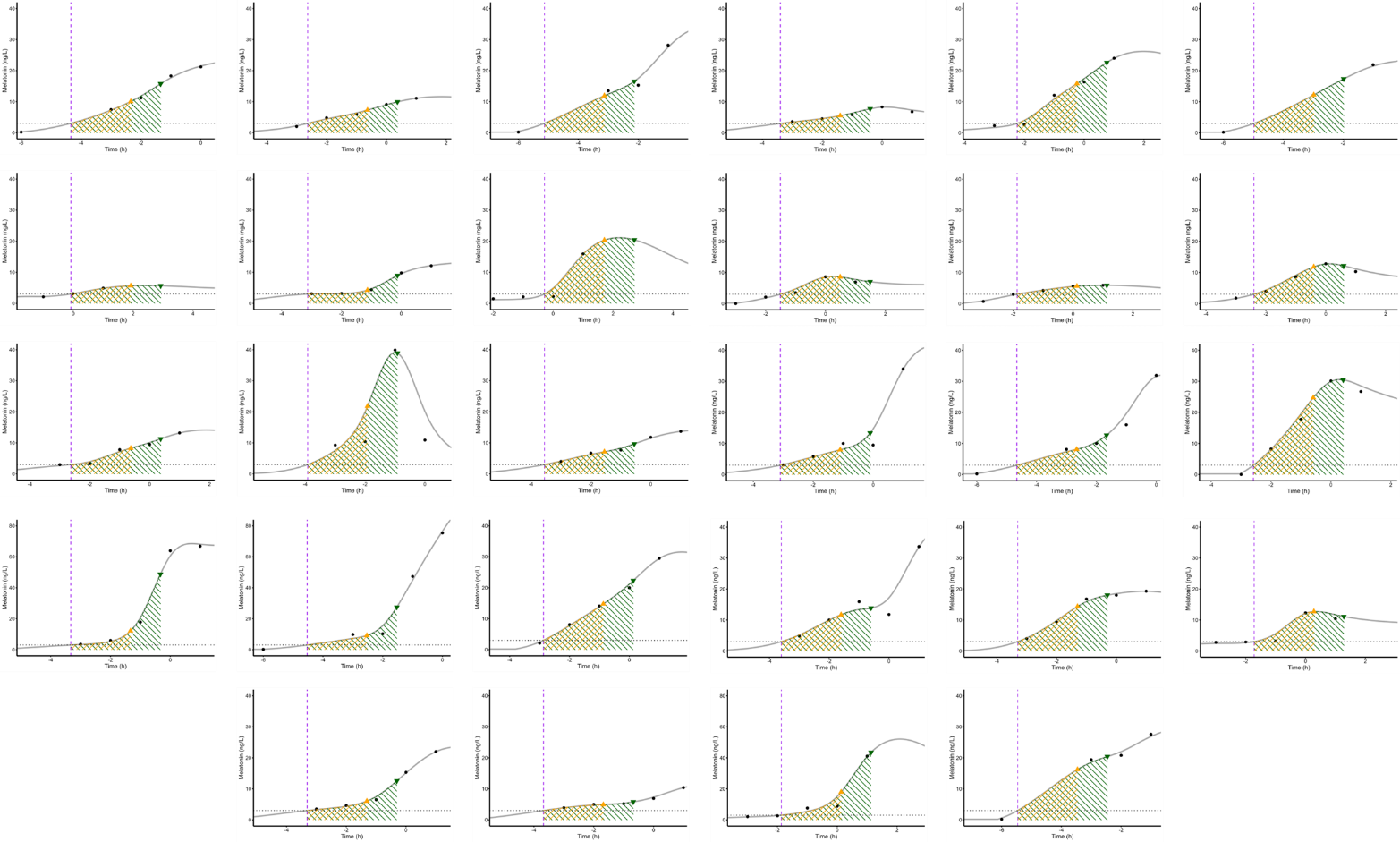

**Figure 2S – MLT secretion curves of plwALS at T6 and derived parameters**

*Legend: purple vertical dash line = DLMO; grey horizontal dot line = MLT levels of 3 pg/ml; black dots = levels of MLT at different time points (at 3, 2, and 1 hours before habitual sleep onset [HSO], at HSO, 1 hour after, and 8 hours after HSO); grey line = line of interpolation of different MLT points; orange triangle = MLT level at DLMO+2 hours (MLT_DLMO2); green triangle = MLT level at DLMO+3 hours (MLT_DLMO3); hatched orange area = Area under the curve between DLMO and DLMO+2; hatched green area = Area under the curve between DLMO and DLMO+3.*
